# Antibody binding to native cytomegalovirus glycoprotein B predicts vaccine efficacy

**DOI:** 10.1101/2020.02.27.20028563

**Authors:** Jennifer A. Jenks, Cody S. Nelson, Hunter K. Roark, Matt Goodwin, Robert F. Pass, David I. Bernstein, Emmanuel B. Walter, Kathryn M. Edwards, Dai Wang, Tong-Ming Fu, Zhiqiang An, Cliburn Chan, Sallie R. Permar

**Author notes:** Corresponding author: Sallie R. Permar. Drs. Chan and Permar contributed equally.

## Abstract

Human cytomegalovirus (HCMV) is the most common infectious cause of congenital disease and post-transplant complications worldwide, yet vaccine development remains hampered by a limited understanding of protective immune responses. We investigated humoral immune correlates of protection against HCMV acquisition elicited by the most efficacious HCMV vaccine tested to-date, soluble glycoprotein B (gB) with MF59 adjuvant (gB/MF59), which achieved ∼50% efficacy in two phase II clinical trials. Protection against primary infection correlated with high magnitude antibody binding to gB expressed on a cell surface, but not to the vaccine antigen. Further, we identified monoclonal antibodies that differentially recognized soluble and cell-associated gB, highlighting structural differences essential for protective immunity. These results indicate the importance of the native, cell-associated gB conformation in future HCMV vaccine design.

**One Sentence Summary:** Partially-effective HCMV gB/MF59 vaccine-elicited IgG binding to cell-associated gB correlates with protection against HCMV

## Introduction

Human cytomegalovirus (HCMV) is a ubiquitous viral pathogen and a leading cause of post-transplant and congenital infections worldwide (*1, 2*). In the United States alone, each year, 40,000 infants are born with congenital HCMV infection, with nearly a third of these children developing permanent hearing loss, brain damage, or neurodevelopmental delays (*3-5*). A vaccine to prevent congenital HCMV has been a “Tier 1 priority” for the U.S. National Academy of Medicine for almost 20 years (*6*), yet no HCMV vaccines are currently licensed for clinical use.

HCMV vaccine development remains hindered in part by limited knowledge of the immune responses that protect against HCMV infection (*7*). Pre-existing immunity to HCMV does not always prevent reinfection nor *in utero* transmission and congenital disease (*8, 9*). Therefore, development of an efficacious HCMV vaccine requires rational vaccine design (*7, 10*). To address this need, we aimed to identify the humoral immune correlates of risk for the most protective HCMV vaccine candidate tested to-date and determine immunologic endpoints important for future vaccines.

Glycoprotein B (gB) is an HCMV envelope glycoprotein highly conserved across the herpesvirus family and is essential for viral entry into all cells, making it an attractive target for vaccination (*11, 12*). Up to 70% of total serum antibodies specific to HCMV are directed against gB (*13*), yet notably, most antibodies generated against gB are non-neutralizing (*14*). The recombinant HCMV gB subunit combined with MF59 adjuvant (Sanofi Pasteur) conferred approximately 50% protection in Phase II clinical trials of HCMV-seronegative adolescent girls (*15*) and postpartum women (*16*). In the gB/MF59 vaccine, the gB sequence was derived from the HCMV Towne strain (genotype 1) and modified by removal of the transmembrane domain and mutagenesis of the furin cleavage site (*17*). Although the gB/MF59 vaccine showed partial protection against primary CMV acquisition, the immune responses that conferred this protection remain unclear.

Previous studies suggested that non-neutralizing antibody responses may have contributed to the partial efficacy of the gB/MF59 vaccine. In HCMV-seronegative transplant recipients, gB/MF59 vaccination reduced the duration of HCMV viremia and antiviral therapy, yet, surprisingly, vaccination did not elicit neutralization of heterologous HCMV strains (*18, 19*). Similarly, in HCMV-seronegative postpartum women, vaccination elicited limited neutralization of the autologous Towne strain and negligible neutralization of heterologous strains (*20*). By contrast, the non-neutralizing function antibody-dependent cell-mediated phagocytosis (ADCP) was measurable in nearly all postpartum vaccinees (*20*). These studies implicated non-neutralizing antibody responses in protection against HCMV primary infection but lacked the statistical power to identify immune correlates of risk for the gB/MF59 vaccine.

In this study, we assessed serum immune responses in the Phase II gB/MF59 clinical trials in postpartum women (ClinicalTrials.gov Identifier NCT00125502) and adolescent girls (NCT00133497) to identify associations between vaccine-elicited humoral immune responses with risk of primary HCMV acquisition. The results of this study will guide HCMV vaccine design and candidate selection.

## Results

### Comparison of vaccine-elicited antibody responses in adolescent (AV) and postpartum vaccinees (PV)

We first compared vaccine-elicited IgG binding responses between the AV and PV cohorts, which demonstrated distinct sera binding profiles (**Table S2**). AV developed higher IgG binding and avidity to full-length gB (**Fig. 1A, B**), gB ectodomain, and Domain I, whereas PV had higher binding to Domain II, Domains I+II, and Antigenic Domain (AD)-1 (**Fig. 1C**). The dominant linear gB epitope IgG response in AV was directed against the cytosolic AD-3 region (**Fig. S1**), as previously reported in PV.(*20*) The cohorts had similar vaccine-elicited IgG binding to gB expressed on epithelial cells (**Fig. S2**), yet AV demonstrated higher magnitude IgG binding to whole HCMV virions (**Fig. S3A, B**).

**Fig. 1.**
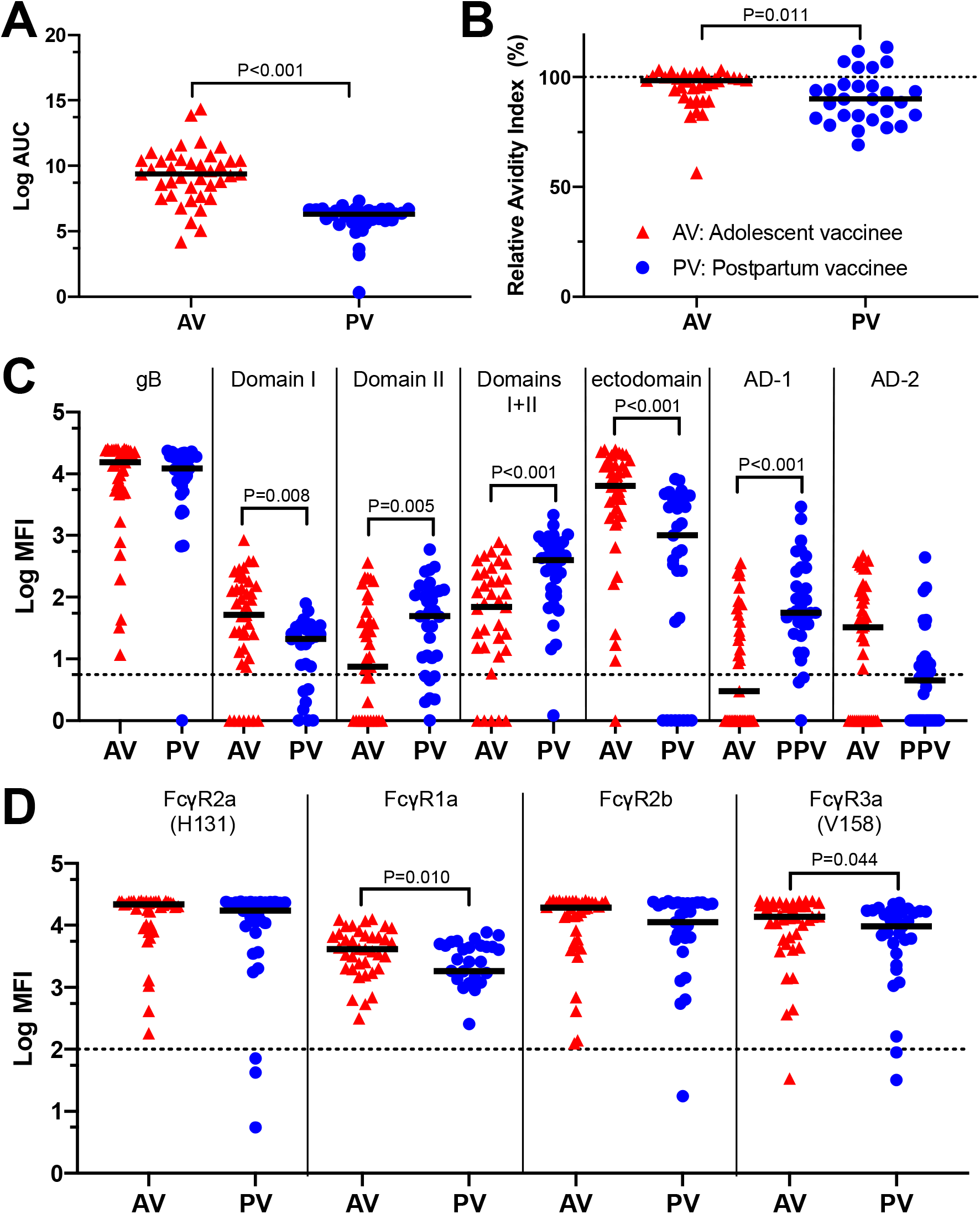
Sera from adolescent and postpartum vaccine cohorts exhibited distinct HCMV and gB-specific IgG binding responses. AV exhibited increased (**A**) IgG binding to full-length gB by enzyme-linked immunosorbent assay (ELISA) (P<0.001) and (**B**) avidity to gB as compared to postpartum vaccinees (P=0.011). Log AUC is the log transformation of the area under the curve (AUC). The relative avidity index is the ratio of antibody binding in the presence or absence of urea. (**C**) AV also had increased magnitude of binding to gB Domain I (P=0.008) and ectodomain (P<0.001), whereas PV had increased magnitude of binding to Domain II (P=0.005), Domains I+II (P<0.001), and gB Antigenic Domain 1 (AD-1. P<0.001) by binding antibody multiplex assay (BAMA). Log MFI is the log transformation of the mean fluorescence intensity (MFI). The dotted line indicates the threshold for positivity, defined by the average of CMV-seronegative samples (n=28) + 2x the standard deviation. (**D**) Cohorts exhibited similar gB-specific IgG binding to Fcγ receptors FcγR2a (H131) and FcγR2b, but AV had increased magnitude gB-specific IgG binding to FcγR1a (P=0.010) and FcγR3a (V158. P=0.044). The dotted line indicates the threshold for positivity of 100 MFI. All cohort comparisons were performed by Mann Whitney U test.

The vaccine-elicited gB-specific IgG Fc region properties and Fc-mediated functions also differed between AV and PV. The cohorts demonstrated similar gB-specific IgG subclass profiles with dominant responses of IgG1 and IgG3 (**Fig. S3C**). Engagement of vaccine-elicited IgG to Fcγ receptors (FcγR) was similar between the cohorts for FcγR2a (H131) and FcγR2b, yet AV demonstrated higher binding than PV to FcγR3a (V158) and FcγR1a (**Fig. 1D**), the high-affinity FcR involved in effector cell activation and ADCP.(*21*) Both cohorts mediated ADCP for heterologous HCMV strains but demonstrated strain-specific differences in phagocytosis (**Fig. 2A**). Consistent with previous reports of poor ADCC responses in PV,(*20*) this response was also poorly elicited in AV (**Fig. S4A**).

**Fig. 2.**
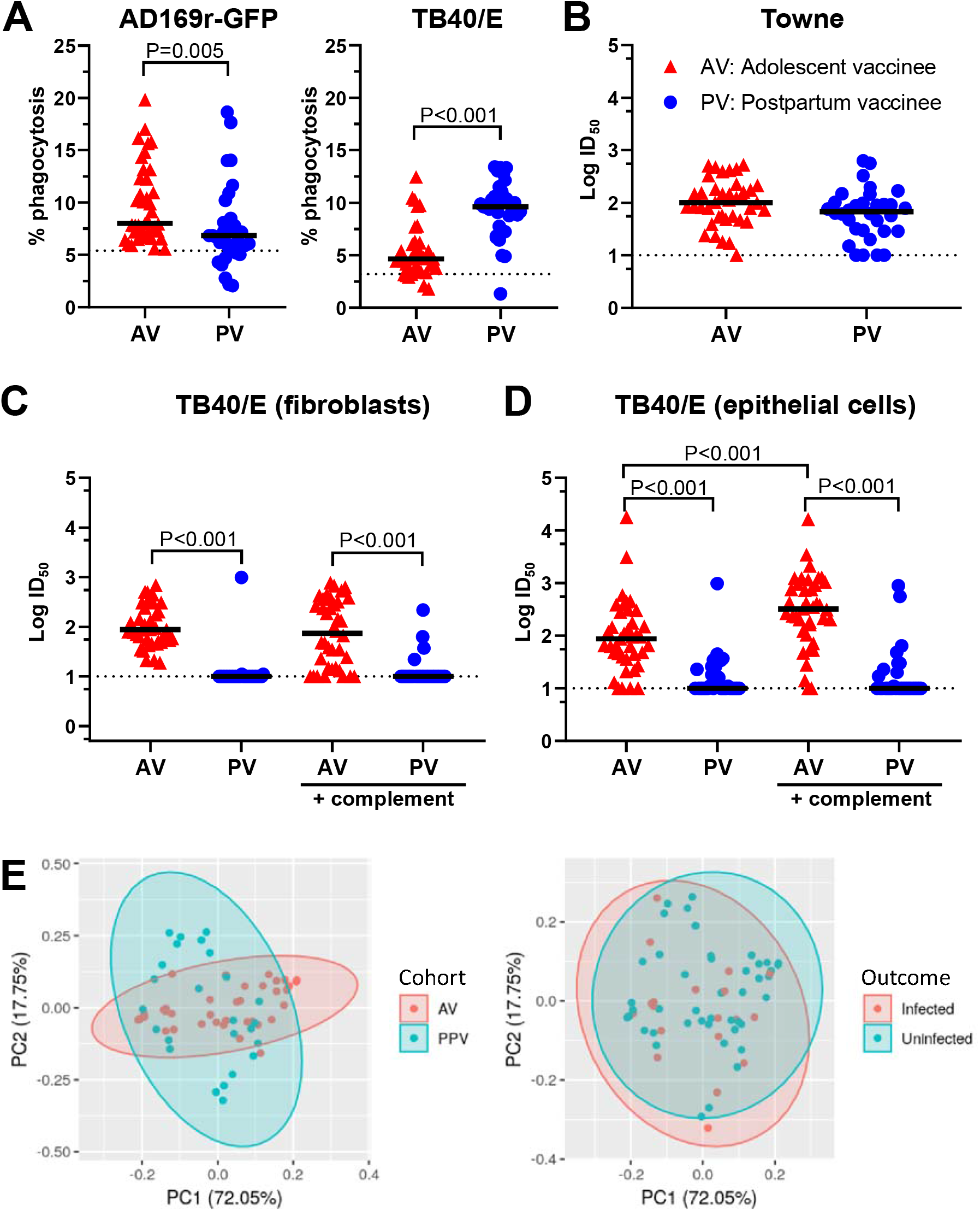
Vaccine-elicited functional antibody responses were distinct between the adolescent and postpartum cohorts. (**A**) Adolescent gB/MF59 vaccinees had higher antibody-dependent cellular phagocytosis (ADCP) of AD169r-GFP strain virus as compared to PV (P=0.005) but lower ADCP of TB40/E strain. The % phagocytosis is reported as the % of total monocyte cells that phagocytosed fluorescently labeled virus. The dotted line indicates the threshold for positivity, as defined by the average of CMV-seronegative samples run simultaneously (n=6) + 2x the standard deviation. (**B**) Autologous neutralization of Towne strain virus was similar in both cohorts. Log ID_50_ is the log-transformed 50% inhibitory concentration in response to the virus strain. For all neutralization assays, the dotted line indicates the threshold of detection (starting sera dilution). (**C**) On both BJ5T-a fibroblasts and (**D**) ARPE epithelial cells, AV exhibited significant neutralization of heterologous TB40/E strain virus (P<0.001), with complement-mediated enhancement of neutralization on epithelial cells (P<0.001), but PV did not demonstrate heterologous neutralization. All cohort comparisons were performed by Mann Whitney U test. (**E**) Principal component analysis (PCA) was used to visualize humoral immunogenicity variables by vaccination cohort and outcome. PC1 and PC2 indicate the principal components along which data have the largest variance.

Vaccination induced similar autologous Towne strain neutralization between the cohorts (**Fig. 2B**), yet, surprisingly, only AV generated robust heterologous virus neutralization responses. AV sera neutralized strains TB40/E and AD169r on both fibroblasts and epithelial cells, with complement-mediated enhancement on epithelial cells, whereas heterologous virus neutralization was negligible in PV (**Fig. 2C, D and Fig. S4B, C**). Interestingly, heterologous virus neutralization inversely correlated with subject age yet was more strongly associated with subject cohort (**Table S3**).

We compared the cohorts by principle component analysis (PCA) across all measured features, excluding heterologous virus neutralization due to limited responses in PV. By PCA, the vaccine immunogenicity profiles of AV and PV were poorly overlapping (**Fig. 2E**). Thus, immune correlate analyses were performed both with cohorts combined and independently.

### Combined immune correlate analysis of the gB/MF59 vaccine

To identify immune correlates of risk of HCMV acquisition, we performed a univariate logistic regression for HCMV infection status in the clinical trial follow-up period. This analysis was performed across 23 measured vaccine-elicited humoral immune responses, excluding heterologous virus neutralization. We adjusted for cohort and performed Benjamini-Hochberg correction with an a priori significance cut-off of <0.2. Regression analysis revealed that only IgG binding to gB-transfected cells correlated with decreased risk of HCMV acquisition (P=0.008, FDR=0.190. **Table 1, Fig. 3A**). This correlate remained significant after adjustment for cohort, timepoint, and subject age (P=0.003, FDR=0.080. See **Table S4**). When assessed in the AV and PV cohorts independently, this immune correlate maintained a trend towards statistical significance (P=0.11, P=0.053, respectively, by Mann Whitney U. **Fig. 3A**). IgG binding to cell-associated gB was not significantly correlated with any other measured humoral response (**Fig. S5**). In a follow-up experiment, we determined whether IgG binding to gB-transfected cells served as a surrogate measure for IgG binding to infected cells. Although infected-cell IgG binding was associated with protection in both cohorts (P=0.036. **Fig. 3B and Fig. S6**), this measure did not correlate with IgG binding magnitude to gB-transfected cells (**Fig. S5**).

**Table 1.** Humoral immune correlate of HCMV acquisition risk of gB/MF59 vaccinees with adjustment for cohort. Logistic regression analysis on 23 measured vaccine-elicited antibody responses, excluding heterologous neutralization responses, revealed that IgG binding to gB-transfected cells was significantly associated with protection against HCMV acquisition (P=0.008, FDR=0.190 by Benjamini-Hochberg correction). FDR indicates the false discovery rate. Log MFI is the log transformation of the mean fluorescence intensity (MFI). Log AUC is the log transformation of the area under the curve (AUC). The % phagocytosis is reported as the % of total monocyte cells that phagocytosed fluorescently labeled virus. Log ID_50_ is the log-transformed 50% inhibitory concentration in response to the virus strain. RAI indicates the relative avidity index, which is calculated as the percentage of antibody binding remaining in the presence of urea.

|  | Assay | p | FDR |
| --- | --- | --- | --- |
| 1 | gB-transfected cell IgG binding (% binding) | **0.008** | **0.190** |
| 2 | Full-length gB IgG binding (LogMFI) | 0.04 | 0.307 |
| 3 | gB Domains I+II IgG binding (LogMFI) | 0.04 | 0.307 |
| 4 | gB ectodomain IgG binding (LogMFI) | 0.058 | 0.334 |
| 5 | ADCP of AD169r-GFP (% phagocytosis) | 0.115 | 0.385 |
| 6 | ADCP of TB40/E (% phagocytosis) | 0.122 | 0.385 |
| 7 | gB Domain II IgG binding (LogMFI) | 0.135 | 0.385 |
| 8 | gB IgG binding (Log AUC) | 0.146 | 0.385 |
| 9 | gB-specific IgG1 subclass (LogMFI) | 0.151 | 0.385 |
| 10 | gB Domain I IgG binding (LogMFI) | 0.169 | 0.389 |
| 11 | Whole virion (TB40/E) IgG Avidity (RAI) | 0.205 | 0.395 |
| 12 | FcγR3a V158 binding to gB-specific IgG (LogMFI) | 0.206 | 0.395 |
| 13 | FcγR1a binding to gB-specific IgG (LogMFI) | 0.294 | 0.507 |
| 14 | FcγR2b binding to gB-specific IgG (LogMFI) | 0.309 | 0.507 |
| 15 | Whole virion (TB40/E) IgG binding (LogAUC) | 0.354 | 0.513 |
| 16 | FcγR2a H131 binding to gB-specific IgG (LogMFI) | 0.357 | 0.513 |
| 17 | gB IgG Avidity (RAI) | 0.451 | 0.610 |
| 18 | gB AD-1 IgG binding (LogMFI) | 0.529 | 0.677 |
| 19 | gB-specific IgG3 subclass (LogMFI) | 0.559 | 0.677 |
| 20 | Neutralization of Towne virus on MRC5 (LogID<sub>50</sub>) | 0.728 | 0.837 |
| 21 | gB-specific IgG2 subclass (LogMFI) | 0.809 | 0.887 |
| 22 | gB-specific IgG4 subclass (LogMFI) | 0.93 | 0.948 |
| 23 | gB AD-2 Site 1 IgG binding (LogMFI) | 0.948 | 0.948 |

**Fig. 3.**
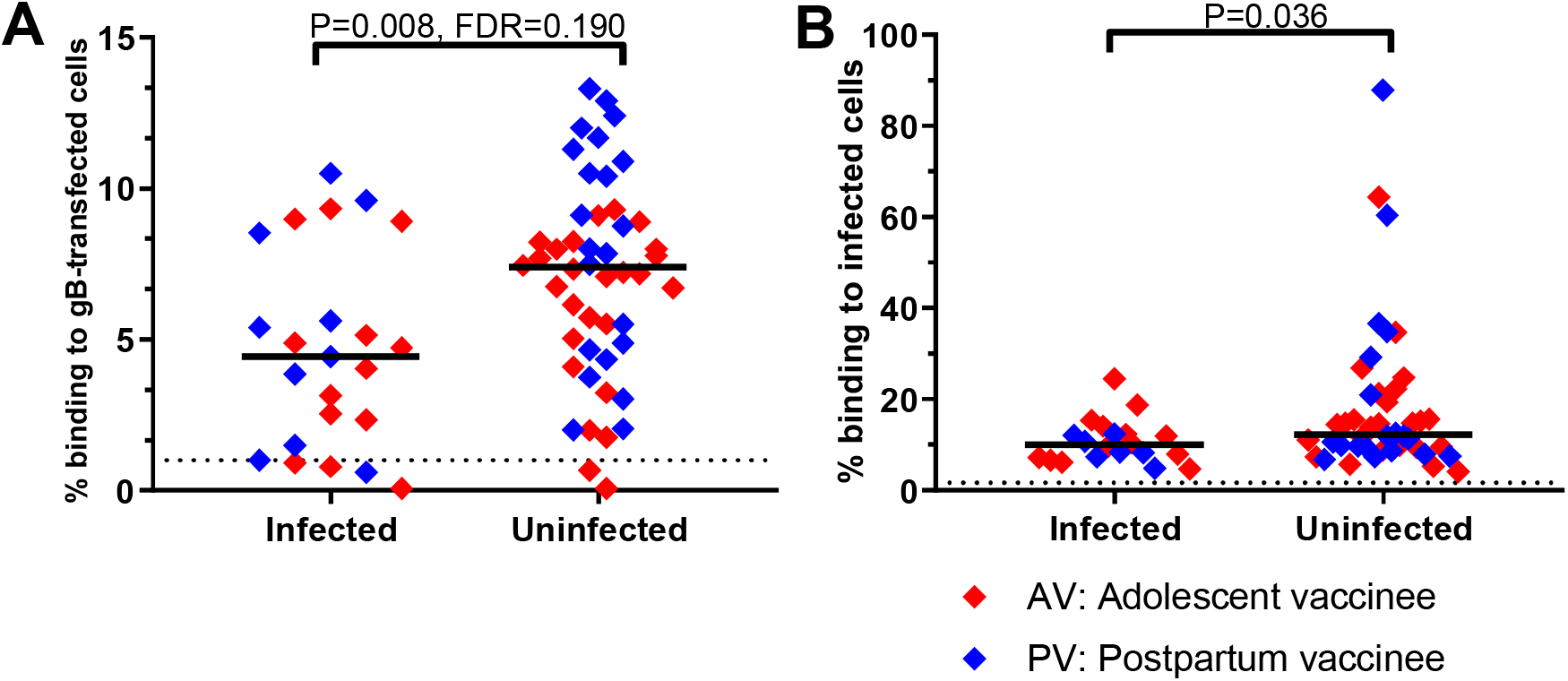
Immune correlates of risk of primary HCMV acquisition elicited by the gB/MF59 vaccine. (**A**) Univariate logistic regression analysis revealed that the magnitude of IgG binding to cell-associated gB was correlated with risk of HCMV primary infection (P=0.008, FDR=0.190 with Benjamini-Hochberg correction). The % binding to gB-transfected cells indicates the % of cells transfected to express gB that are bound by IgG in vaccinee sera. The dotted line indicates the threshold for positivity, as defined by the average of CMV-seronegative samples run simultaneously (n=10) + 2x the standard deviation. (**B**) gB/MF59 vaccinees that acquired HCMV had lower magnitude sera IgG binding to fibroblasts infected with AD169r-GFP HCMV strain as compared to uninfected vaccinees (P=0.036) by Mann Whitney U test. The % binding to infected cells indicates the % of HCMV-infected cells that are bound by IgG in vaccinee sera. The dotted line indicates the threshold for positivity, as defined by the average of CMV-seronegative samples run simultaneously (n=8) + 2x the standard deviation.

To validate IgG binding to gB-transfected cells as a measure distinct from binding to soluble gB, we compared the binding of gB-specific monoclonal antibodies (mAbs) to these gB forms. A panel of gB-specific mAbs isolated from three naturally-infected individuals demonstrated similar magnitude binding to soluble gB. However, they had a bimodal distribution of IgG binding strength to gB-transfected cells (**Fig. 4**), suggesting distinct epitope recognition of cell-associated and soluble gB.

**Fig. 4.**
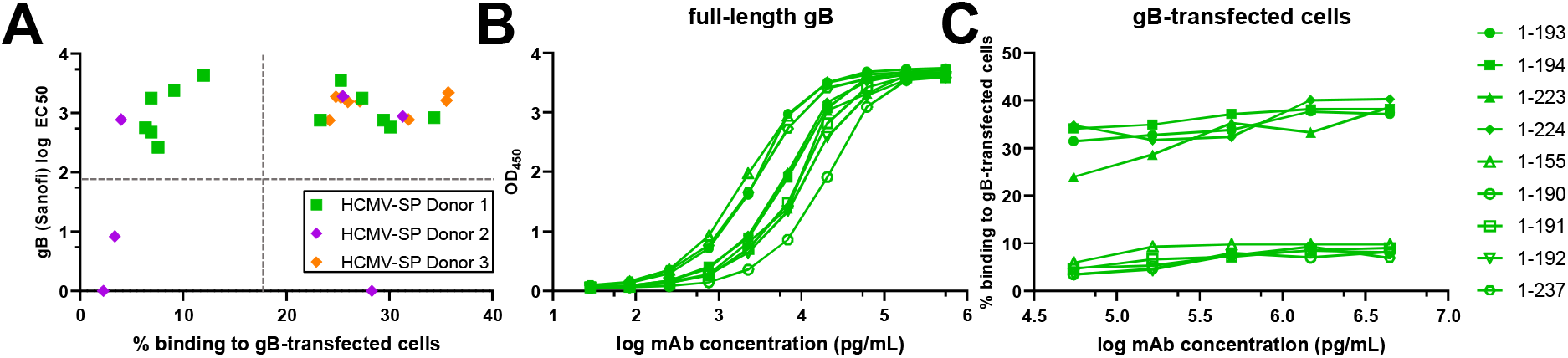
gB-specific mAbs isolated from naturally HCMV-infected individuals that bind with similar strength to full-length gB yet differentially recognize cell-associated gB. (**A**) 26 gB-specific mAbs were recombinantly produced from gB-specific memory B cells isolated from three naturally infected HCMV-seropositive (SP) individuals. mAb binding to full-length gB was measured by ELISA, and binding to gB-transfected cells was measured by flow cytometry at 5 µg/mL. (**B**) mAbs isolated from one donor were run in serial dilution. They demonstrated similar binding kinetics to full-length gB by ELISA. (**C**) However, these mAbs had bimodal binding kinetics to gB-transfected HEK293T cells.

Finally, we built a predictive model of HCMV infection outcomes using a subset of measured immune responses. We removed collinear variables using a variance inflation factor (VIF)<10 criterion, then selected independent predictors of infection outcome by LASSO regression, in which cross-validation was used to select the inverse regularization strength parameter (**Fig. 5A**). To evaluate the robustness of feature selection, we iteratively resampled the dataset with replacement and repeated LASSO regression 1000 times (**Fig. 5B**). In >95% of LASSO samplings, IgG binding to gB-transfected and infected cells, as well as autologous Towne strain neutralization, had non-zero correlations (**Fig. 5B**). Using only LASSO-selected variables, we calculated an ROC curve that was moderately predictive of HCMV infection in gB/MF59 vaccines (AUC=0.78±0.08 for out-of-sample testing. **Fig. 5C, D**).

**Fig. 5.**
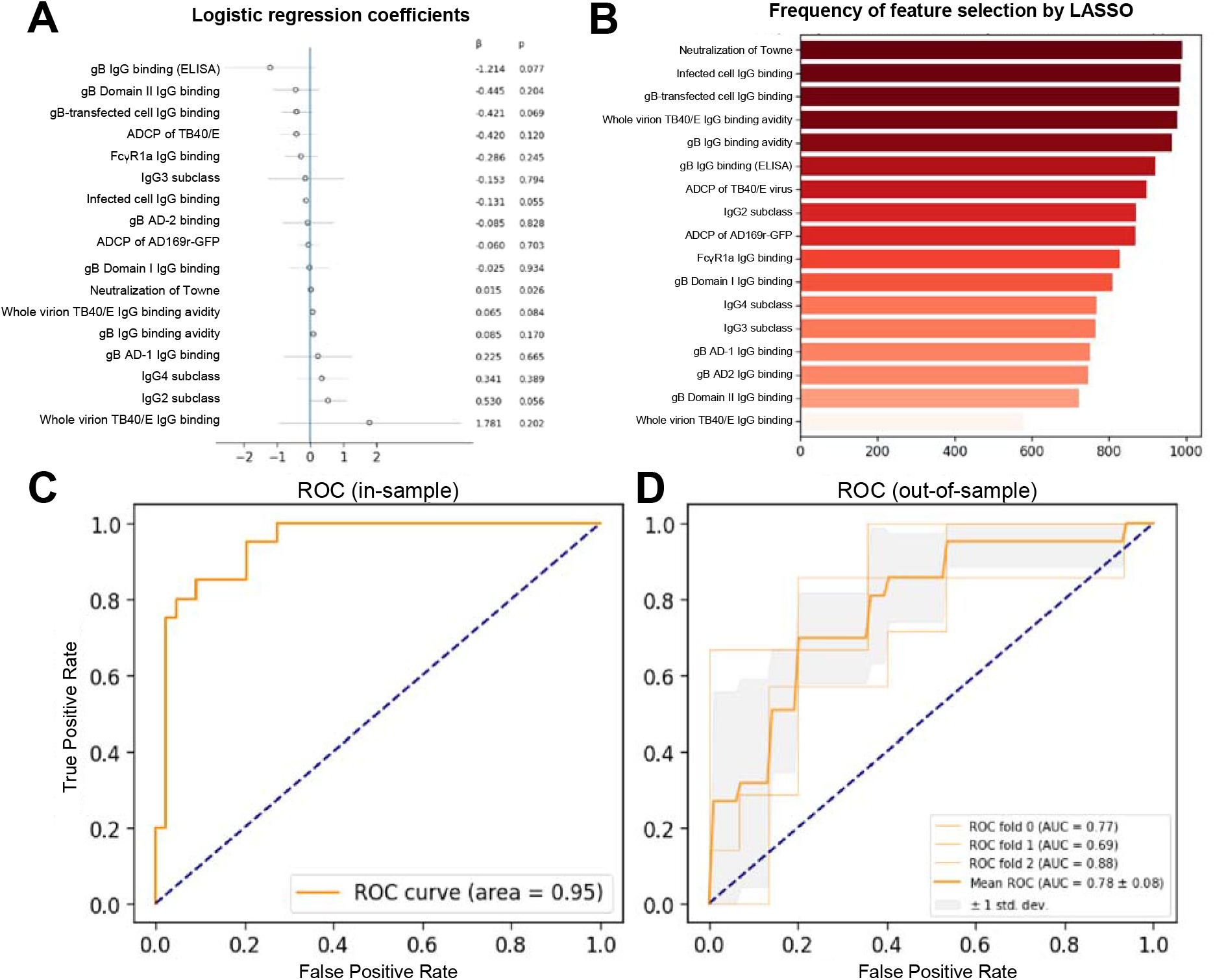
In-sample and out-of-sample receiver operator characteristic curves for a penalized multiple logistic regression model using humoral immunogenicity features to predict outcome of combined vaccine cohorts. (**A**) After exclusion of heterologous neutralization results and collinear variables (variance inflation factor (VIF)>10), a LASSO (least absolute shrinkage and selection operator) regression analysis was performed to generate the feature coefficients. (**B**) LASSO techniques were applied to 1000 bootstrapped samples to evaluate robustness of feature selection. The stability selection plot indicates the number of times that each feature had non-zero coefficients from performing LASSO on each bootstrapped sample. Receiver operator characteristic (ROC) curves and the corresponding area under the curve (AUC) were calculated, with 1 indicating perfect discriminatory value and 0.5 or less indicating no discriminatory value. ROC using in-sample data for the cohorts combined (**C**) and for out-of-sample data (**D**) are shown. Out-of-sample results were obtained by 3-fold stratified cross-validation, including a nested cross-validation within each fold for selection of the inverse regularization strength hyperparameter.

## Discussion

This study reveals an immune correlate of risk of HCMV acquisition for the most efficacious HCMV vaccine candidate trialed to-date. We compared 32 gB/MF59 vaccine-elicited humoral immune responses in adolescent and postpartum cohorts and identified differences in vaccine-elicited immune responses between cohorts, in particular, heterologous virus neutralization. The immune correlate analysis in the combined cohort identified that serum IgG binding to cell-associated gB, but not to the soluble vaccine immunogen or HCMV-neutralizing responses, was associated with protection. Moreover, we identified gB-specific mAbs that differentially bind to cell-associated conformations of gB despite equivalent binding to soluble gB. These findings indicate that antibodies recognize distinct epitopes on cell-associated and soluble gB and that cell surface epitopes of gB may more closely represent those present on viral particles, wherein gB is in its native conformation as a fusogenic protein for HCMV. Therefore, cell-associated conformation of gB may be important for eliciting protective gB-specific antibodies, a major implication for future HCMV gB vaccine design.

HCMV gB mediates membrane fusion for viral entry and is thought to exist in two distinct conformations: prefusion and postfusion. The trimeric postfusion state has been crystalized (*22, 23*), but the structure of the prefusion form, which has fusogenic function on viral particles and is therefore potentially more relevant to the induction of protective antibodies (*24*), remains to be determined. Cell-associated gB may be dynamic and toggle between prefusion and postfusion states (*25*). However, notably, the soluble gB used in the gB/MF59 vaccine studies is primarily monomeric and may not adequately mimic either state (*26*). We theorize that soluble gB may exhibit structural flexibility and form trimeric complexes that enable exposure of relevant binding epitopes on cell-associated gB in some vaccinees. Our findings suggest that future gB-targeted vaccines should direct immune responses against the cell-associated gB conformation, through the use of platforms such as viral vectors, mRNA, or DNA encoding gB (*10, 27*).

In the adolescent and postpartum populations, the gB/MF59 vaccine conferred similar rates of protection against HCMV primary infection yet demonstrated differential vaccine immunogenicity. Interestingly, subject age inversely correlated with heterologous virus neutralization responses. The HPV-16/18 AS04-adjuvanted vaccine is also known to elicit higher anti-HPV-16 and -18 antibody titers in adolescent girls compared to young women (*28, 29*), suggesting that there are distinct immune landscapes in these age groups. Yet, variance in neutralization was better explained by subject cohort than by subject age, indicating that factors other than age, such as recent pregnancy, contribute to vaccine immunogenicity. Our results identify differences in vaccine-elicited immune responses between these cohorts that requires further study and consideration in future HCMV vaccine trial design.

The function mediated by cell-associated gB IgG engagement as an immune correlate of risk of HCMV acquisition remains to be identified. In our study, the neutralizing and non-neutralizing functions ADCC and ADCP did not predict HCMV acquisition risk and did not strongly correlate with IgG binding to cell-associated gB. Furthermore, heterologous virus neutralization was elicited in AV, but not PV, yet this response did not confer additional vaccine efficacy. Thus, our study indicates that although neutralization is the most common immunologic endpoint used in viral vaccine development, this may not be a comprehensive measure to assess experimental HCMV vaccines.

Additional immune responses warrant further study for their roles in protection against HCMV infection. HCMV spreads directly from cell-to-cell, and antibody inhibition may contribute to rapid local HCMV clearance from the mucosa (*30*). gB is known to have five highly homologous genotypes, and whether the gB/MF59 vaccine conferred genotype-specific protection remains unclear (*31, 32*). This study focused on vaccine immunogenicity at single timepoint, but the timing and duration of anti-CMV responses are important in other contexts, including congenital transmission (*33*). Immune recognition of other HCMV glycoproteins, such as the HCMV gH/gL/gO trimer and pentameric complex that generate potently neutralizing responses, may also contribute to protection (*34*). Furthermore, although this study has implicated IgG binding to cell-associated gB in defense against primary infection, its role against secondary infection or reactivation has yet to be studied.

There are limitations to the statistical methods applied. Our univariate regression analysis indicated that IgG binding to cell-associated gB is positively associated with protection, but it cannot identify a specific cutpoint. Furthermore, the ROC curves generated from a select subset of measured immune responses had only moderate predictive ability. However, as a hypothesis-generating study, these results provide future directions for assays that can measure a threshold for protection against HCMV infection.

Our study indicates that vaccine-elicited IgG binding to gB-transfected cells is an important immunologic endpoint for vaccine development and evaluation and a surrogate for an antibody mediated antiviral function. Immunologic endpoints with established relationships to HCMV acquisition will aid in vaccine evaluation in preclinical and early phase clinical trials and should continue to be evaluated in ongoing late phase HCMV vaccine development.

## Data Availability

The complete immunogenicity datasets and reproducible Jupyter notebooks for all analysis performed will be later provided in the Supplementary Materials.

## Acknowledgements

The authors would like to acknowledge Sanofi Pasteur, Merck, and Trellis Biosciences for the generous gift of research materials and the Duke Vaccine and Treatment Evaluation Units for their assistance in study coordination. Furthermore, we would like to thank the subjects who participated in these clinical trials and clinical and research staff at the participating sites.

## Author contributions

J.A.J., C.S.N., C.C., and S.R.P. conceptualized the study; J.A.J. performed all assays with assistance from C.S.N., H.K.R., and M.G.; C.C. and J.A.J. performed all data analysis; C.S.N. and S.R.P. acquired funding; R.F.P., D.I.B., E.B.W., and K.M.E. acquired and provided clinical samples; T.M.F. and Z.A. provided HCMV monoclonal antibodies; S.R.P. provided resources and performed project administration; S.R.P. supervised the study; J.A.J. wrote the original draft; J.A.J., C.S.N., R.F.P., D.I.B., E.B.W., K.M.E., D.W., T.M.F., Z.A., C.C. and S.R.P. reviewed and edited the final manuscript.

## Data and materials availabilities

The complete immunogenicity datasets and reproducible Jupyter notebooks for all analysis performed are provided in the Supplementary Materials.

